# Seroepidemiological Study of Novel Corona Virus (CoVID-19) in Tehran, Iran

**DOI:** 10.1101/2021.01.18.20248911

**Authors:** Zeinab Tabanejad, Sorena Darvish, Zeinab Borjian Boroujeni, Seyed Saeed Asadi, Morteza Mesri, Omid Raiesi, Muhammad Ibrahim Getso, Mahdi Zareei

## Abstract

A novel severe acute respiratory syndrome coronavirus 2 (SARS-CoV-2) has now spread to all countries of the world, including Iran. Although new anti-coronavirus antibodies in patients may be identified by immunological methods with sufficient sensitivity and specificity, the conclusive diagnosis of the disease is by the molecular RT-PCR process. We used a population-based seroepidemiological survey to quantify the proportion of the exposed population with SARS-CoV-2 antibodies and evaluated whether the antibodies are a marker of total or partial immunity to the proportion of the population that remains susceptible to the virus. This cross-sectional study was conducted to investigate the seroprevalence of CoVID-19 in Tehran, the capital of Iran, between April and end of October 2020. Specimens of clotted and heparinized blood (2ml) were collected from the patients. The serum and plasma were separated and stored at − 80LJ°C until use. We examined serum anti-SARS-CoV-2 IgG and IgM antibodies from 1375 in-patients admitted to our hospitals using ELISA kits. In total, 1375 participants were enrolled in this study, and SARS-CoV-2 antibodies were detected using IgM-IgG antibody assay in 291 patients. Among the seropositive patients studied, 187 were men (64.3%), and 104 were women (35.7%) (P<0.05). The mean age of the patients was 49±8.4 years; the majority (27%) were in age group 31-40 years. Also, the lowest frequency of cases was reported in the age group of 1-10 years (P <0.05). We determined the seroprevalence of SARS-CoV-2 for IgM or IgG antibodies to be 21.2%. Diabetes mellitus was the most common underlying disease among SARS-CoV-2 patients [P=0.05; Odd Ratio=1.61(0.90-2.91)]. Conventional serological assays in SARS-CoV-2 cases, such as the enzyme-linked immunoassay (ELISA) for specific IgM and IgG antibodies, have a high-throughput advantage and minimize false-negative rates that occur with the RT-PCR method. This study determined the seroprevalence of SARS-CoV-2 antibodies to be 21%. Control of diabetes among other cases factors shall play important role in management and control of COVID-19.

## Introduction

Since emergence of the novel severe acute respiratory syndrome coronavirus 2 (SARS-CoV-2) in Wuhan, China, on 12 December 2019, it spread quickly across the world and developed into a pandemic[1]. Iran is one of the Middle East countries most affected by the COVID-19 pandemic. The first report of SARS-CoV-2 infection in Iran was reported in the city of Qom on 19 February 2020[2]. COVID-19 disease presentation is highly variable; it ranges from asymptomatic to mild respiratory pneumonia and even severe acute respiratory syndrome (SARS). Asymptomatic or subclinical infections are one of the most important public health challenges of COVID-19 because they spread the infection and remain undetected in the community. Therefore, vigilant control measures are necessary at every stage of the COVID-19 epidemic to avoid spread and resurgence of cases. Among other variables, age has been identified as a risk factor for a more severe course of the disease; younger people tend to have moderate or even asymptomatic presentations and therefore play important role in spreading the infection[3]. Accordingly, one study revealed that 51% of the confirmed cases, including 10 ship crew and 308 passengers, were asymptomatic [4]. Although the definitive diagnosis of the disease requires real-time polymerase chain reaction (RT-PCR), immunological and serological tests can detect coronavirus antibodies with appropriate sensitivity and specificity in patients within the range of presentation (asymptomatic, mild, moderate, and severe) [5]. Combined detection of IgM and IgG is of great value to improve the sensitivity of early diagnosis of COVID-19 [6]. These immunoglobulins, which usually appear in the blood of patients 8 to 10 days after the initial symptoms, can be promptly detected by immunological and serological tests with acceptable sensitivity and specificity, and thus can be an important tool for early detection of asymptomatic carriers in an attempt to control spread of the infection [7-9]. Tehran is the capital and the largest city in the country. It is the most densely populated and reportedly has the largest number of coronavirus cases. However, the incidence rate per 100,000 people is not as high as that of less densely populated provinces of Semnan, Qom, Markazi, Yazd, Mazandaran, Qazvin, Guilan, Alborz, and Isfahan. Thus, public health awareness campaigns in large and densely populated cities - where most sensitive jobs are located - is of economic and social importance because increasing cases in these cities can impact on other cities in the country. [2]. The proportion of the population that has SARS-CoV-2 antibodies can be quantified by a population-based seroepidemiological survey. The survey can provide information on proportion of the exposed population; the population that remains susceptible to the virus; and whether the antibodies are a marker of total or partial immunity [10]. This study aimed to determine the seroprevalence of SARS-CoV-2 antibodies, and the risk factors associated with SARS-CoV-2 infection in Tehran to provide valid decision-grounds for healthcare professionals to effectively prevent, control, and treat the infection.

## Materials and methods

### Study design and samples

This cross-sectional study was conducted at Valiasr, Sajad and Ghaem hospitals, Tehran, the capital of Iran from April to the end of October 2020 to investigate the seroprevalence of COVID-19. The hospitals are the referral centers for patients within and outside Tehran. With their written consent, we included 1375 COVID-19-suspected patients according to the Helsinki Declaration. Blood samples of patients were collected according to the hospital’s infection control guidelines along with adequate personal protective equipment. During sample collection, we administered questionnaires to collected patient’s information, such as age, gender, occupation, underlying disease, and other essential information from the patient’s records.

### ELISA assay

Both clotted and heparinized blood (2ml each) were collected from the patients. The serum and plasma were separated and stored at − 80□°C until use. We examined for anti-SARS-CoV-2 IgG and IgM antibodies in serum samples from 1375 participants using ELISA kits (IDEAL TASHKHIS Co, IR. Iran). For detection of IgM, 100 µL of diluted serum (1:50) was added into the 96-well microplate (coated with N protein) and then incubated for 30 min at 37°C. After washing, 100 µL of enzyme conjugate was added into the wells and then incubated for another 30 minutes at 37°C. Following the second wash cycle, 100 µL of substrate was added into the wells and incubated for 15 minutes under 25°C. Finally, a stop solution was added to the wells to terminate the reaction. The optical density (OD) of each well was determined by a microplate reader at 450 nm within 30 minutes. For detection of IgG, the dilution factor was changed (1:100). According to the protocol, the sensitivity of the test was about 61% for IgM antibodies between days 7 and 14 of infection and 82% for IgG antibodies between days 11 and 15. The performance of the kit was confirmed on 20 samples of PCR-negative volunteers (Negative control) and 20 patients with definitive coronavirus (PCR positive) (Positive control).

### Statistical analyses

We analyzed the obtained data using SPSS Ver.22.0 software employing statistical tests such as Chi-Square, and Fisher’s Exact Test. P-value <0.05 was considered significant.

## Results

In total, 1375 participants were enrolled in this study, and SARS-CoV-2 was detected in 291 patients using IgM-IgG antibody tests. Out of 1375 participants studied, 954 were men (69.4%), and 421 women (30.6%) (P<0.05). The mean age of the patients was 49±8.4 years; the majority (27%) fall within 31-40 years. Also, the lowest cases were reported in the age group of 1-10 years (2%) (P <0.05). Among all participants, 291 patients (21.2%) were positive for either IgM or IgG antibodies, indicating past or present infection (P <0.05). Among 291 seropositive patients, 187 (64.3%) were men and 104 (35.7%) were women (P<0.05). The summary of our serological findings are shown in **Table 1**. A significant difference was observed between age over 60 years (as a risk factor) and other age groups [P <0.05; Odd Ratio = 2.72 (1.7-4.35)]. **(Table 2)**.

**Table 1.** Seroprevalence of new coronavirus patients by gender

|  | **IgM or IgG** |  | **Total** |
| --- | --- | --- | --- |
|  | **Positive** | **Negative** |  |
| **Male** | 187 (13.6%) | 767 (55.8%) | **954 (69.4%)** |
| **Female** | 104 (7.6%) | 317 (23%) | **421 (30.6%)** |
| **Total** | 291 (21.2%) | 1084 (78.8%) | **1375 (100)** |
| **P=0.03, df=1** |  |  |  |

**Table 2.** Seroprevalence of new coronavirus patients by age groups

|  | **IgM or IgG** |  | **Total** |
| --- | --- | --- | --- |
|  | **Positive** | **Negative** |  |
| 0-10 | 6 (0.5%) | 16 (1.2%) | **22 (1.6%)** |
| 11-20 | 15 (1.1%) | 76 (5.5%) | **91(6.6%)** |
| 21-30 | 51 (3.7%) | 286 (20.8%) | **337 (24.5%)** |
| 31-40 | 75 (5.5%) | 296 (21.5%) | **371 (27%)** |
| 41-50 | 72 (5.2%) | 259 (18.8%) | **331 (24.1%)** |
| 51-60 | 40 (2.9%) | 104 (7.6%) | **144 (10.5%)** |
| >60 | 32 (2.3%) | 47 (3.4%) | **79 (5.7%)** |
| **Total** | 291 (21.2%) | 1084 (78.8%) | **1375(100)** |
| **P=0.0001, df=6** |  |  |  |

Clinical and laboratory data of the patients were collected and analyzed. Fifty-seven patients (4%) had underlying diseases including diabetes, cardiovascular diseases, autoimmune disorders, respiratory diseases, etc. Diabetes mellitus was the most common underlying disease among SARS-CoV-2 patients [P=0.05; Odd Ratio=1.61(0.90-2.91)]. Twenty-eight percent of the participants were government employees, and the rest were self-employed, or unemployed. A comparison of the monthly incidence of new cases of coronavirus in government employees and other occupations is shown in **Figure 1**. Daily new cases and death from COVID-19 in Iran at 6 December 2020 have been shown in **Figure 2**. Among the seropositive patients, 66 (22.7%) were in the group of government employees and 225 (77.3%) were among the others [P <0.05; Odd Ratio = 0.79 (0.6-0.98)] **(Figure 3)**.

**Fig. 1.**
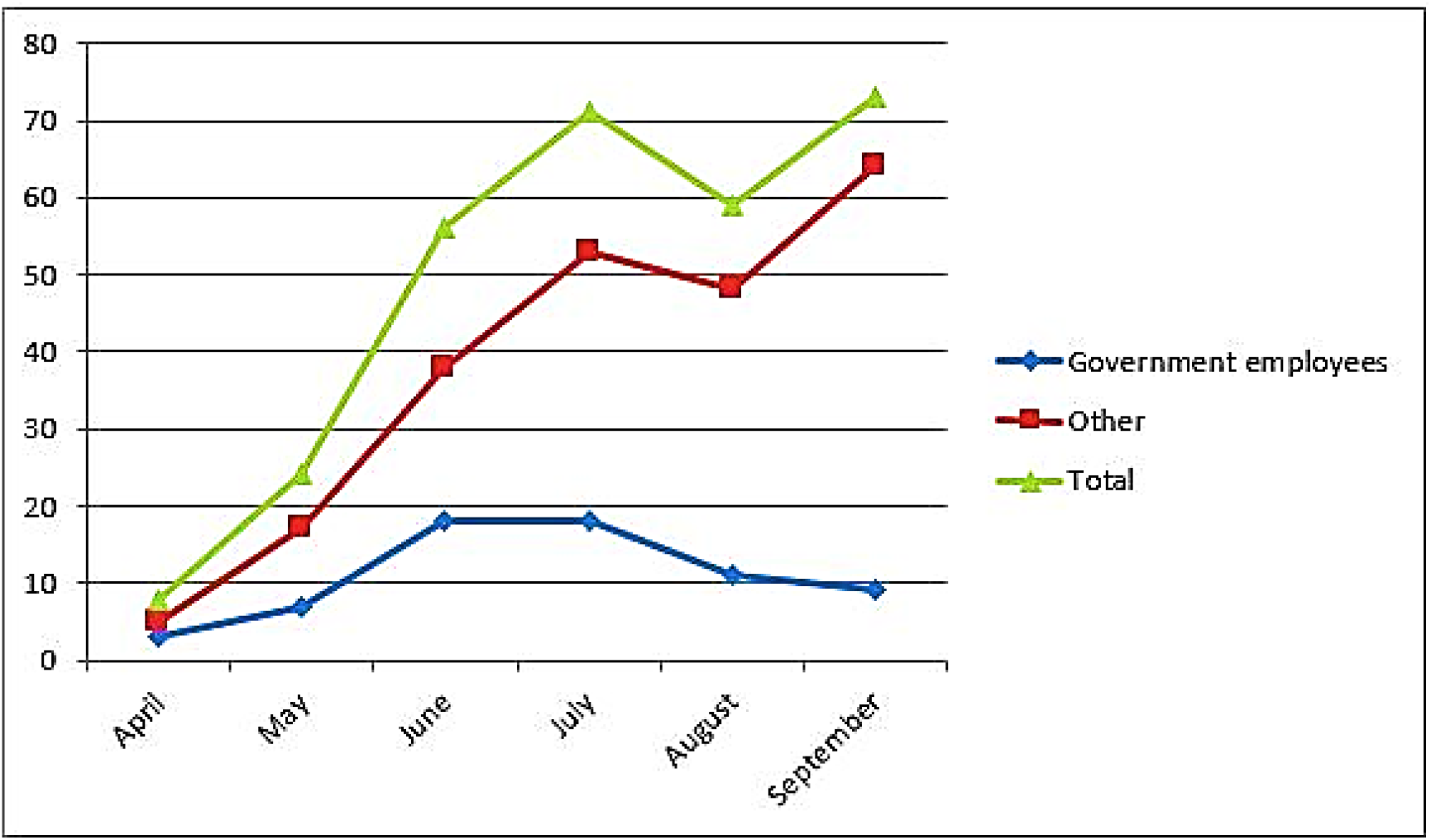
Comparison of monthly incidence of new cases between government employees and other occupations.

**Fig. 2.**
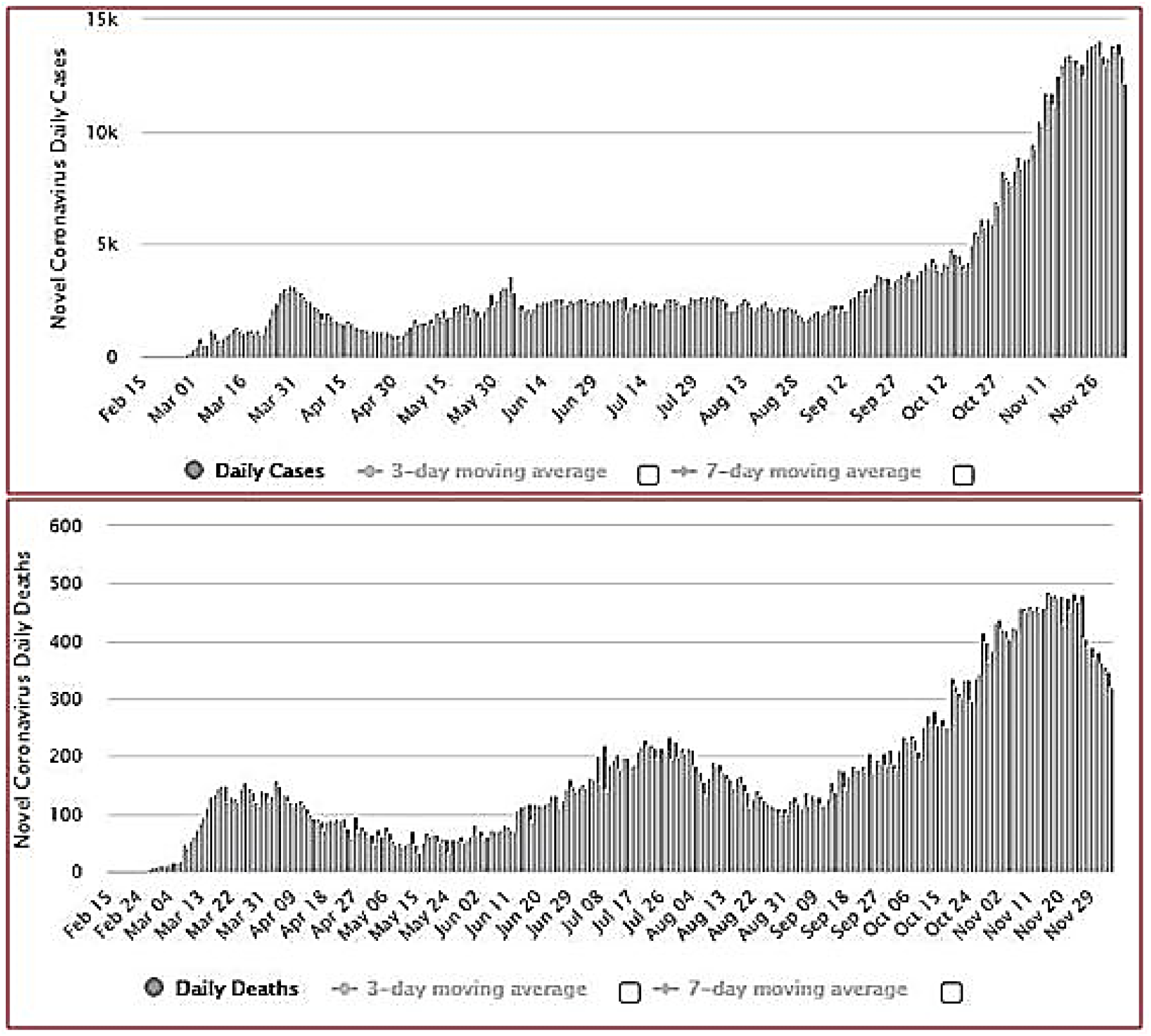
Coronavirus Cases in Iran. Daily New Cases (Upper) and Daily New Deaths (Lower).

**Fig. 3.**
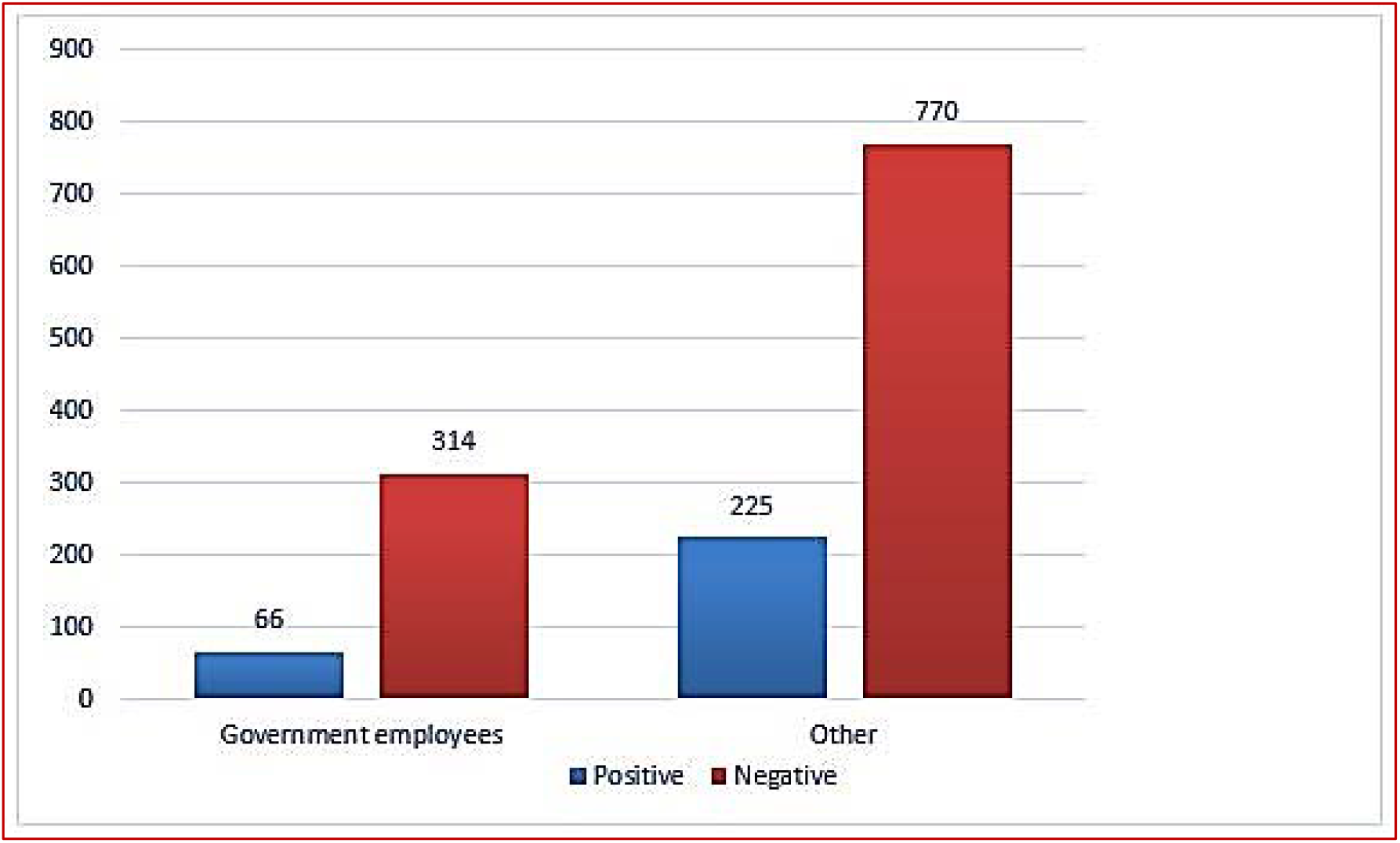
Seroprevalence of new coronavirus patients by type of occupation

## Discussion

COVID-19 emerged as a global threat, affecting 73,809,570 people worldwide and causing about 1,642,000 deaths as of 16 December, 2020. According to WHO reports, the overall mortality rate for COVID-19 was 2.3%. With more than 1,123,000 confirmed cases and more than 52,000 deaths by December 16, Iran remains one of severely affected countries in the Middle East. Individuals with undocumented illnesses, either as asymptomatic and do not seek medical treatment, or those that are infectious throughout the incubation phase pose the greatest risk of transmission[3]. The proportion of asymptomatic infections reported in different studies varies greatly, ranging from 4% to 41%[11]. This finding reinforces the importance of aggressive measures to identify, treat, or isolate individuals with confirmed SARS-CoV-2 infection and their contacts to halt the spread of the epidemic. Despite the definitive diagnosis of the disease by molecular RT-PCR, immunological and serological tests can detect coronavirus antibodies (immunoglobulins) in patients at different stages of the infection with acceptable sensitivity and specificity. A serological survey is a powerful tool to determine the spread of infectious diseases, particularly in the presence of asymptomatic cases or incomplete ascertainment of those with symptoms[10]. From the current study, the seroprevalence of SARS-CoV-2 antibodies in Tehran was 21.2%. Our result corroborates that reported in the study conducted at Guilan province[4]. However, the result is much higher than the seroprevalence estimated in California, USA, which was between 2.49% and 4.16%[12]. Besides, if we want to compare the outcome of serological methods with that of molecular tests (RT-PCR) in Iran, the results of molecular tests have reported a lower prevalence of 12% - according to the report of the Ministry of Health of Iran by the end of October, 4,599,554 tests were performed, of which 545,286 were positive [13]. The reasons for this discrepancy include the ease of access to antibody testing, low cost, and the desire of clients for cheaper and more accessible methods and false-negative results in molecular testing [14, 15]. Statistically, a comparison of monthly incidence from the beginning of May to the end of October, the general frequency of cases was ascending (P <0.05), but the frequency of monthly incidence in government employees after attaining a peak in July and August, it started decreasing and among the unemployed or self-employed, it was still rising **(Figure 1)**. The mandate to follow health protocols among government employees can be one of the reasons for this downward trend after August [P<0.05; Odd Ratio=0.79(0.6-0.98)]. These results indicate successful commitment by government employees to adhere to health protocols through a unified and coordinated management system. However, the lack access by non-employed and self-employed to the government’s imposed and coordinated management system, impacted the prevalence of the disease; making it higher. In our study, men were more affected than women (P <0.05) **(Table 1)**, and this may be related to their exposure. Due to the special culture of Iranians, men are more likely than women to work outside the home, more at risk than women and even the mortality rate in women is lower than men. In our study, the mean age of the patients was 49±8.4 years; the majority (27%) were at 31-40 years. Also, the lowest case frequency was reported in the age group of 1-10 years (2%) (P <0.05). These results are in line with the results of a study conducted in Spain, and one of the possible causes of these results is that the age groups 21 to 50 years are active occupational groups, and the incidence of these occupational groups is higher than other age groups. However, the intragroup comparison of people over 60 years old showed that out of 79 patients, 32 (41%) had coronavirus infection, compared to other groups with high relative risk of infection [P <0.05; Odd Ratio = 2.72 (1.7-4.35)] **(Table 2)**. This pronounces the needs for more effective planning specifically tailored to the elderly. According to the statistical results, failure to observe health protocols increases the prevalence of the disease. General measures to prevent the spread of COVID-19 include effective quarantine and isolation of infected cases, close monitoring of primary contacts, travel restrictions, and personal safety measures. Besides, extension of health protocols to specific occupational groups and individuals with underlying conditions should be done by the authorities [16-19].

## Conclusions

This study determined the seroprevalence of SARS-CoV-2 antibodies of 21% in Tehran. We also demonstrated that the conventional serological assays, such as the enzyme-linked immunoassay (ELISA) for specific IgM and IgG antibodies, have a high-throughput advantage and minimizes the false-negative cases that occur with the RT-PCR method. It is hoped that the current findings could provide valid decision-grounds for healthcare professionals to effectively prevent, control, and treat SARS-CoV-2 infections amid the current pandemic. Control of diabetes among other factors shall play important role in management and control of COVID-19.

## Data Availability

The datasets used and/or analysed during this study are available from the corresponding author on reasonable request.

## Acknowledgments

Sincere gratitude to all staff, professors, and students at Tehran University of Medical Sciences, Iran University of Medical Sciences, Shahid Beheshti University of Medical Sciences, as well as Sajad, Ghaem, and Valiasr Hospitals.

## Ethical approval

The work was approved by the ethic committee of the Iran University of Medical Science (IUMS), Tehran, Iran (no. IR.IUMS.FMD.REC.1399.485). Informed consent was obtained from all individual participants included in the study.

